# Heterogeneity of recurrences in rectal cancer: application of population models facilitates personalized medicine

**DOI:** 10.1101/2021.09.02.21263020

**Authors:** Iris D. Nagtegaal, Carlijn van de Water, Dyogo Borst, Corrie A.M. Marijnen, Cornelis J.H. van de Velde, Niek Hugen, Eelke Jongejans

## Abstract

Due to heterogeneity in presentation and outcome, patients with metastatic disease cannot be considered a single group. The timing, location and combinations of recurrences determine the feasibility of treatment of the individual patient in an era in which the options for local and systemic treatment have expanded. Studies investigating this complexity are hampered by the lack of both large cohorts and adequate methods.

In a well-defined cohort of rectal cancer patients from a randomized clinical trial, with long standardized follow-up, we applied spatial projection models derived from population ecology to overcome the complexity problem. We describe the recurrence patterns in detail and performed stochastic simulation experiments resulting in 1.5 million evaluable patients. The risk of subsequent recurrences was dependent on the presentation of the first recurrent event and decreased with increasing recurrence-free interval. The risk of local recurrence for the median patient (65.8 years, pT3 adenocarcinoma) was threefold increased after the development of rare metastases. The risk of development of rare metastases was increased after the development of other extrahepatic metastases.

Our cross-disciplinary approach delivers insights allowing for the development of personalized strategies for (local) treatment of recurrent disease, as well as for surveillance strategies that may potentially impact large patient cohorts. In this proofof-principle study we demonstrate the feasibility of spatial projection models for cancer research.

## Introduction

Changes in colorectal cancer treatment have significantly improved survival for patients with metastatic disease over the past years. Early detection of metastases due to improved imaging techniques(1-3) in combination with more effective forms of local and systemic therapy(4-7) have significantly impacted survival of patients with metastatic disease(8).

It has become clear that patients with recurrent disease cannot be regarded as a homogenous group. Recurrences include both local recurrences as well as distant metastases. We previously showed that there are considerable outcome differences between synchronous and metachronous metastases, with the latter having a poor overall response rate to first line systemic therapy(9). Population-based studies show that there is variation in outcome dependent on the location of metastatic disease, with decreased outcomes reported for patients with peritoneal metastases(10) compared with liver and lung metastases. Smaller studies indicate that rare metastatic locations, such as brain(11, 12) and bone(13) metastases are also associated with poor outcomes. Differences in local treatment options and response to systemic treatment(10) can only partially explain these differences. Further insight into relationships between patterns of recurrence, patterns of progression and final outcome are crucial to tailor treatment for recurrent disease. In order to predict the success of local treatment of recurrent disease, we need to improve our understanding of sequential events: which patients are indeed adequately treated and which factors determine the recurrence risk for patients after treatment of the first recurrence. Insight into patterns of metastatic spread, defined by location, timing and sequence of events will be of utmost importance. Currently, these multimodal analyses are hampered by the lack of sufficiently large cohorts with detailed and standardized follow-up and by the restrictions of standard statistical approaches. The number of potential variables and combinations requires a novel approach. We therefore propose a spatial projection model, based on mathematical models derived from the field of population ecology(14). In that particular field these models are used to study the impact of environmental factors on the persistence, growth, and spread of populations of a wide variety of species. The simulations based on population models enable i) a systematic analysis of the changes in outcome influenced by certain metastatic events, and ii) risk assessment on the level of individual patients.

In this study, we analyze a well-defined cohort of rectal cancer patients, treated within the framework of a large multicenter randomized trial(15), with standardized surveillance and follow-up. We will explore the possibilities of the use of population models in cancer progression, by combining traditional statistics with transition matrix modelling, to study combinations of metastases and timing of events in more detail. As proof of principle, we will demonstrate the validity and value of our approach and its possibilities for the improvement of personalized treatment of patients with recurrent rectal cancer.

## Methods

### Patient population

In the TME trial, patients with a resectable adenocarcinoma of the rectum were randomly assigned to preoperative radiotherapy with 5×5 Gy followed by total mesorectal excision (TME) within a week or to TME alone. The design of the trial and the main outcomes have been reported previously(15). Central and local ethics committee approval for the study was obtained as well as informed consent from all included patients.

Patients underwent clinical examination every 3 months during the first year after surgery and annually thereafter. Local recurrence was defined as evidence of tumor within the pelvic or perineal area. Criteria for distant recurrence involved tumor growth in any other area, including the colostomy site or inguinal region. The Data Center at the Department of Surgery of the Leiden University Medical Center in the Netherlands managed primary and follow-up data on case report forms that were collected by participating hospitals and general practitioners, until July 15, 2010. In addition, a search was performed at the national digital pathology archive (PALGA: Pathologisch – Anatomisch Landelijk Geautomatiseerd Archief)(16), at 01/04/2017 under search number LZV 2017-50 to retrieve additional details of all follow-up histology and possibly autopsy information to complete the information on disease progression.

### Statistics for parameterizing a spatial population model

In order to model recurrence dynamics, we first restructured the database by subdividing each patient’s medical history into trimesters (i.e. 3-month periods) starting with the day of TME surgery. For each trimester transition we noted which recurrences were known at the beginning of the transition and which new ones had been discovered meanwhile. Once a recurrence was discovered, the affected organ was set to ‘with metastasis’ for the rest of the patient’s life, irrespective of any (local) treatment. This allowed us to analyze nine probabilities: i) the risk of death during the first trimester after surgery, and during subsequent trimesters, ii) the risk of recurrence-related death, iii) the risk of death due to unrelated (conditional on surviving metastatic disease) causes. The remaining six probabilities were (conditional on survival) the chances that whether during a trimester for the first time a recurrence is discovered at a new location: iv) local recurrence, v) liver metastases, vi) lung metastases, vii) distant lymph node metastases, viii) peritoneal metastases or ix) rare metastases. For each of these nine response variables we fitted logistic regression model with the following additive explanatory variables (without interactions): age at time of surgery, sex, invasion depth of the primary tumor (classes ‘pT1-2’, ‘pT3’ or ‘pT4’), number of regional lymph node metastases, radiotherapy treatment, time since surgery, and whether or not there had been any recurrences already at each of the six locations (local recurrence, liver, lung, distant lymph nodes, peritoneum, rare metastases). Rare metastases include metastases in ovaries (n = 5), urine bladder, kidney (n = 4), adrenal gland (n = 4) thyroid, uterus, heart, stomach, breast (n = 2), sometimes in combinations. For the mortality model for the first trimester after surgery the latter six explanatory variables were reduced to one: whether or not any synchronous metastases were discovered at the time of surgery. All continuous variables were normalized (mean=0, standard deviation=1) prior to the analyses. We applied a full model averaging approach using the dredge function of the MuMIn R Package.

### Stochastic simulations of the patient population and of ‘median’ patients

The averaged regression models of these nine possibilities were then used in two sets of stochastic simulations to explore a) metastatic patterns and b) sensitivity of metastatic outcome to initial conditions and model parameters (i.e. to what extent does a small change in a parameter result in changes in outcome). The simulations started at the moment of surgery: each trimester mortality and new metastases randomly occurred, with probabilities depending on model predictions for the current situation. Each simulation was run until the death of the simulated patient, after which the order and timing of metastatic spread was recorded as well as the cause of death. In the first set of simulations, the initial conditions (i.e. the age, sex, and treatment of the patient, as well as the characteristics of the initial tumor and synchronous metastases) matched those of the 1,452 patients in turn, each repeated a thousand times, resulting in nearly 1.5 million simulations.

For the second set of simulations we created eight patient profiles that all had the cohort’s median age at the time of surgery (65.8 years) and a pT3 adenocarcinoma, but who systematically differed in a full-factorial way in sex, radiotherapy treatment, and number of lymph nodes affected (0 or 3; the latter being the median number of positive lymph nodes in stage III patients). Each of these eight patient profiles were then exposed to seven initial patterns of metastatic disease: no metastases, or one metastasis at one of the six locations. These simulations started either immediately after initial surgery or two years later (assuming survival and no further metastatic spread until that point). Each of these 8×7×2=112 scenarios were simulated a thousand times.

### Statistical effects of metastases at a single location

To illustrate how single metastases contribute to metastatic spread we also fitted a separate set of regression models. Assuming patient survival over the coming quarter year, we analyzed the chance of a new metastases being discovered at a particular organ, given a metastasis already being present at a particular other organ but not at any of the other organs. Fitted regression models did include the same patient characteristics (e.g. age and sex) as before. These regression models were then used to calculate (for an average female patient) the probabilities of new metastases appearing once single organs are colonized.

### Conventional statistics

The χ^2^ test was used to compare proportions in demographic, clinical and pathological characteristics by type of recurrence. In survival analyses overall survival was defined as the interval between the date of surgery until the date of death or until last follow-up. Patients who were alive at the end of follow-up were censored in survival analyses. Overall survival curves were generated according to the Kaplan-Meier method and equality of distributions was compared with log-rank testing. Conditional cox regression testing was used to retrieve additional information about the relevance of prognostic factors. All tests of significance were two-tailed and differences at P-values of <0.05 were considered significant.

## Results

Of the 1,530 patients that were included in the Dutch cohort of the trial, we excluded the patients who did not undergo resection of the primary tumor (N=76), as well as one patient who died at the day of surgery and one patient who was lost to follow-up. Out of the remaining 1,452 patients (observation cohort), 495 patients presented with metastatic disease, either at the moment of diagnosis (synchronous metastases, TNM stage IV, n=93, i.e. 18.8% of the 495) or during follow-up. Local recurrence was diagnosed in 130 patients during follow-up. The clinical data of these patients are summarized in table 1.

**Table 1.** Overview of the clinical data categorized according to the location of the first recurrence. Percentages are given per row, except for the number of locations of recurrence, where percentages per column are shown. Patients without recurrent disease are not taken into account for statistical analyses. * Rare metastases are grouped together in the category other, data per location are given in supplemental table 1. P-values are derived from anova testing (age) and chi-square testing (other variables). RT: preoperative radiotherapy (5×5 Gy). TNM: Tumor Node Metastasis classification.

| First site of recurrence |  |  |  |  |  |  |  |  | p-values |
| --- | --- | --- | --- | --- | --- | --- | --- | --- | --- |
|  |  | None | Local recurrence | Liver metastasis | Lung metastases | Distant lymph nodes | Peritoneal metastases | Other metastases* |  |
|  | No. of cases | 926 | 91 | 257 | 92 | 31 | 29 | 28 |  |
| age | Mean (years) | 64.7 | 65.0 | 62.7 | 64.2 | 61.8 | 65.7 | 64.8 | 0.41 |
| gender | Female | 65.1% | 6.6% | 15.4% | 5.9% | 2.7% | 2.7% | 2.0% | 0.43 |
|  | Male | 62.9% | 6.0% | 19.0% | 6.6% | 1.8% | 1.6% | 1.7% |  |
| treatment | RT | 66.8% | 3.0% | 17.2% | 6.9% | 2.3% | 1.8% | 2.1% | 0.64 |
|  | No RT | 60.6% | 9.5% | 18.2% | 5.8% | 1.9% | 2.2% | 0.4% |  |
| Distance from the anal verge | < 5 cm | 60.5% | 7.8% | 15.5% | 8.0% | 4.4% | 1.8% | 2.0% | 0.011 |
|  | 5-10 cm | 63.3% | 7.0% | 18.4% | 6.3% | 1.5% | 1.9% | 1.7% |  |
|  | 10-15 cm | 67.6% | 3.6% | 19.1% | 4.6% | 0.5% | 2.4% | 2.2% |  |
| TNM stage | I | 88.6% | 1.6% | 5.0% | 2.7% | 0.9% | 0.5% | 0.7% | <0.001 |
|  | II | 73.7% | 6.0% | 11.3% | 5.0% | 0.8% | 1.0% | 2.3% |  |
|  | III | 46.5% | 11.3% | 23.1% | 10.9% | 3.4% | 1.9% | 2.9% |  |
|  | IV | 0.0% | 1.1% | 74.2% | 3.2% | 6.5% | 14.0% | 1.1% |  |
| Tumor type | Adenoca. | 64.3% | 6.1% | 18.3% | 6.0% | 1.8% | 1.7% | 1.8% | 0.015 |
|  | Mucinous ca | 59.2% | 7.9% | 12.5% | 9.2% | 3.9% | 4.6% | 2.6% |  |
| No. of locations with recurrences | 0 | 100% | - | - | - | - | - | - | < 0.001 |
|  | 1 |  | 33.3% | 61.8% | 57.6% | 69.0% | 53.6% | 74.1% |  |
|  | 2 |  | 30.3% | 23.7% | 28.3% | 24.1% | 28.6% | 18.5% |  |
|  | 3 |  | 18.0% | 10.0% | 12.0% | 6.9% | 10.7% | 7.4% |  |
|  | 4 |  | 15.7% | 3.6% | 2.2% | - | 7.1% | - |  |
|  | 5 |  | 2.2% | 0.8% | - | - | - | - |  |

### First recurrence location

The majority of patients presented with liver metastases as first recurrence (48.8%), followed by lung metastases (17.5%) and local recurrences (17.3%). Location of first recurrence was influenced by preoperative radiotherapy, which decreased the local recurrence risk (p < 0.001), by tumor type (with less liver metastases for mucinous carcinomas, p = 0.015) and by the location of the primary tumor. The low rectal cancers (0-5 cm from the anal verge) showed more lung metastases, less liver and less peritoneal metastases. The increased number of distant lymph node metastases in the low rectal cancers was caused by the involvement of the inguinal lymph nodes. There was an overall increased risk of recurrence with increasing stage, from 11.4% in stage I patients to 53.5% in stage III patients (p < 0.001).

The location of the first recurrence significantly impacted survival (figure 1A), with a median survival after recurrence of 28.3 (95%CI 23.7-33.0) and 29.1 months (95%CI 21.3-36.9) for liver and lung metastases, respectively. Patients with distant lymph node metastases had a median survival of 14.7 months (95%CI 9.7-19.7) and patients with peritoneal metastases 12.0 months (95%CI 6.5-17.5). Patients with a local recurrence had the worst outcome with a median overall survival of 7.7 months (95%CI 5.5-9.8). There was no difference in survival between the randomization groups.

**Figure 1:**
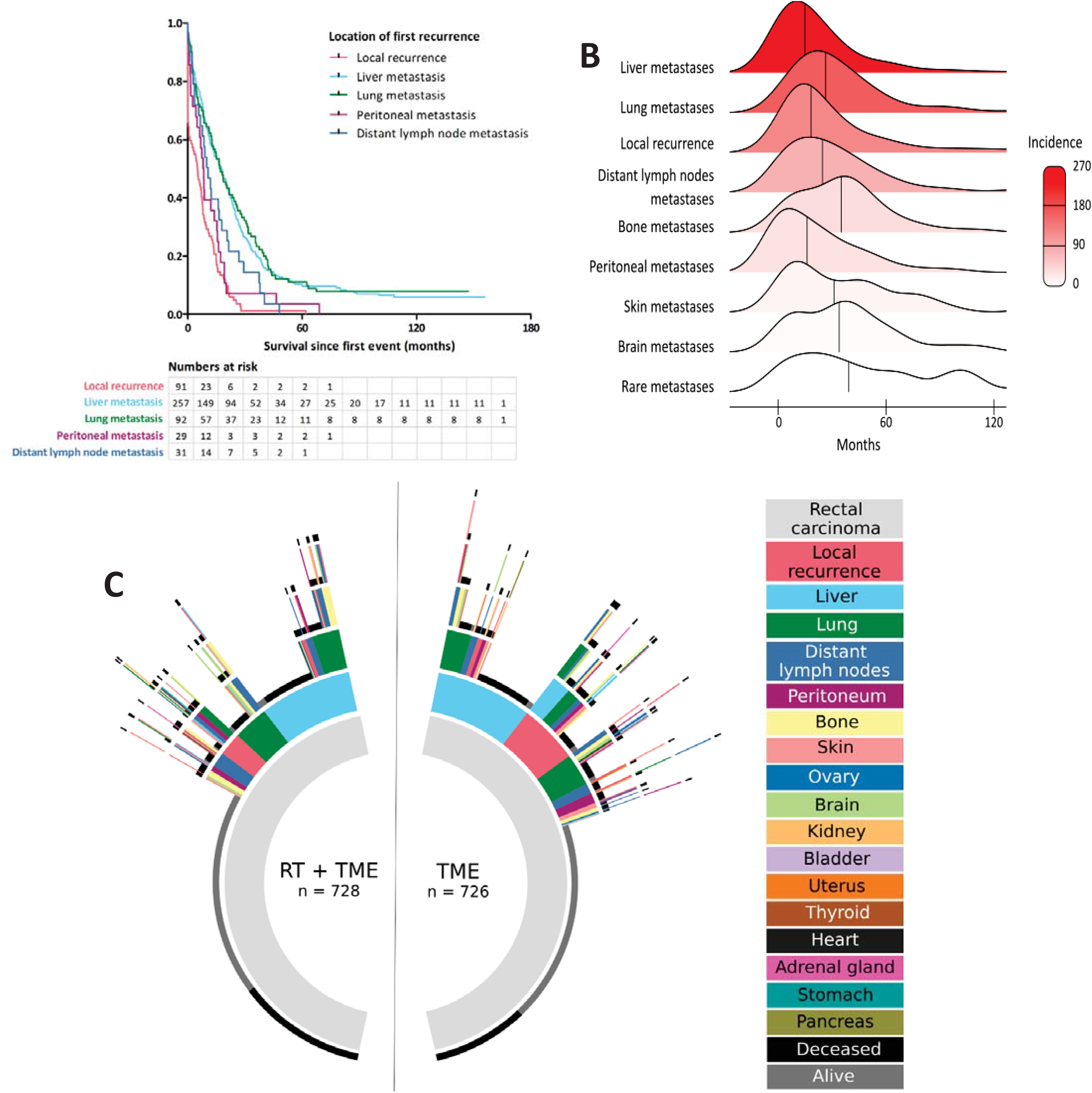
Metastatic patterns in the observation cohort. **A. Impact of first metastatic location on outcome.** Kaplan-Meier curve illustrating survival since detection of first metastasis: There is a significantly better survival probability for patients with lung and liver metastases, compared to other metastases (p < 0.001). **B. Timing of (metachronous) metastases**. Stacked density plot of the whole cohort illustrating the rapid development of liver metastases and the relative late development of rare metastases (p < 0.001). **C. Sequence of recurrences according to the type of treatment**. The inside rings indicate the earlier events. Preoperative radiotherapy (RT) decreases the risk of local recurrence (in yellow), in this group less diversity in metastatic patterns is observed (63 versus 80 patterns). TME: total mesorectal excision, indicates the surgery only group.

### Timing of recurrence

Of the 92 synchronous metastases, the majority was present in the liver (74.2%, stage IV, table 1), followed by peritoneal metastases (14%). The timing of metachronous recurrence differed according to their location (p < 0.001, figure 1B), with rapid development of liver metastases (50% of the liver metastases developed within 17 months, 95%CI 14.3-19.8 months), compared to the other metastatic locations. Timing of metastases was not different between patients who were treated with either preoperative radiotherapy or surgery only, except for skin metastases (after radiotherapy: median interval of 19.9 months, 95% CI 12.0-27.8 months, versus surgery: median interval 49.7 months, 95% CI 42.3-57.1 months). Most skin metastases were present in the abdominal wall (14/28) and perineum (8/28). There was no difference in distribution over the randomization arms, although the majority of cases developed in patients with surgery-only treatment (19/28).

For local recurrences, time to diagnosis was significantly shorter in the surgery-only group (median 16.1 months, 95%CI 13.6-18.6) compared to the radiotherapy group (32.1 months, 95%CI 28.4-35.9). Local recurrence was the first event in 91 patients and a later event in 39 patients. Patients with local recurrence as a first event developed metastases in 67.6% of cases. The timing of those subsequent distant recurrences is different from the overall distribution, with earlier development of skin and distant lymph node metastases, and later development of brain metastases.

### Sequence of recurrences

In figure 1c sequential recurrence patterns for the individual patients are depicted per randomization arm. In the radiotherapy group 63 different patterns can be distinguished, versus 80 patterns in the surgery only group. Single-organ recurrences were present in 56.7% of cases (table 1). The liver was the most common site for single-organ distant recurrence (52.5%). Patients with lung or peritoneal metastases presented more often with other metastatic locations during follow-up. Patients with a single metastatic location had a better prognosis compared to patients with 2 or more metastatic locations (26.1 months (95%CI 22.1-30.1) versus 19.1 months (95%CI 15.3-22.9), p = 0.028). This difference in outcome is mainly caused by the overrepresentation of patients with liver metastases in the group with a single metastatic location, as is evident by conditional forward cox regression analysis (with both the location of the first metastasis as well as the number of metastases).

### Model validation for the patient population

To enable stochastic simulation of metastatic spread, nine (model-averaged) regression models were fitted (figure 2a, see details in Appendix) and were used to run near 1.5 million stochastic simulations, we checked the validity of our simplified model (not including interactions) by evaluating the modelled life spans and metastatic patterns. Simulated male and female patients reached an average age of 68.1 (SD 10.6) and 69.4 (SD 11.3) respectively when dying of metastatic disease, without metastases they reached an average age-at-death of 77.8 (SD 8.2) and 80.0 (SD 8.7) years. The general pattern of metastatic spread was very comparable between the patient and the simulated cohort. Timing (figure 2b), sequence and distribution of recurrences according to location of metastasis was similar between the observation data and the simulations (figure 2c). Notable differences are that some synchronous metastases were already detected days before surgery, while the simulations set these at t=0 due to their strict trimester time step. On the other end of the time spectrum, the simulations showed a longer tail of new metastases appearing long after initial surgery. This reflects the fact that the simulations ran until the death of the simulated patient, while the trial had a very decent, but limited duration of follow-up (median 11.6 years), leading to right-censored data. The validity of the simulations was also confirmed by comparing the observed order of metastatic spread (Fig. 1b), with those in the simulations (Fig. 2b). As the simulations also ran longer than the follow-up of the clinical trial, there were more simulated than observed patients with metastases in at least 1 organ (42.0% vs 32.9%, respectively). As there were a thousand-fold times more simulations than observed patients, the diversity of unique sequences of metastatic spread was higher in the simulations and included rare cases of spread to (nearly) all organs.

**Figure 2.**
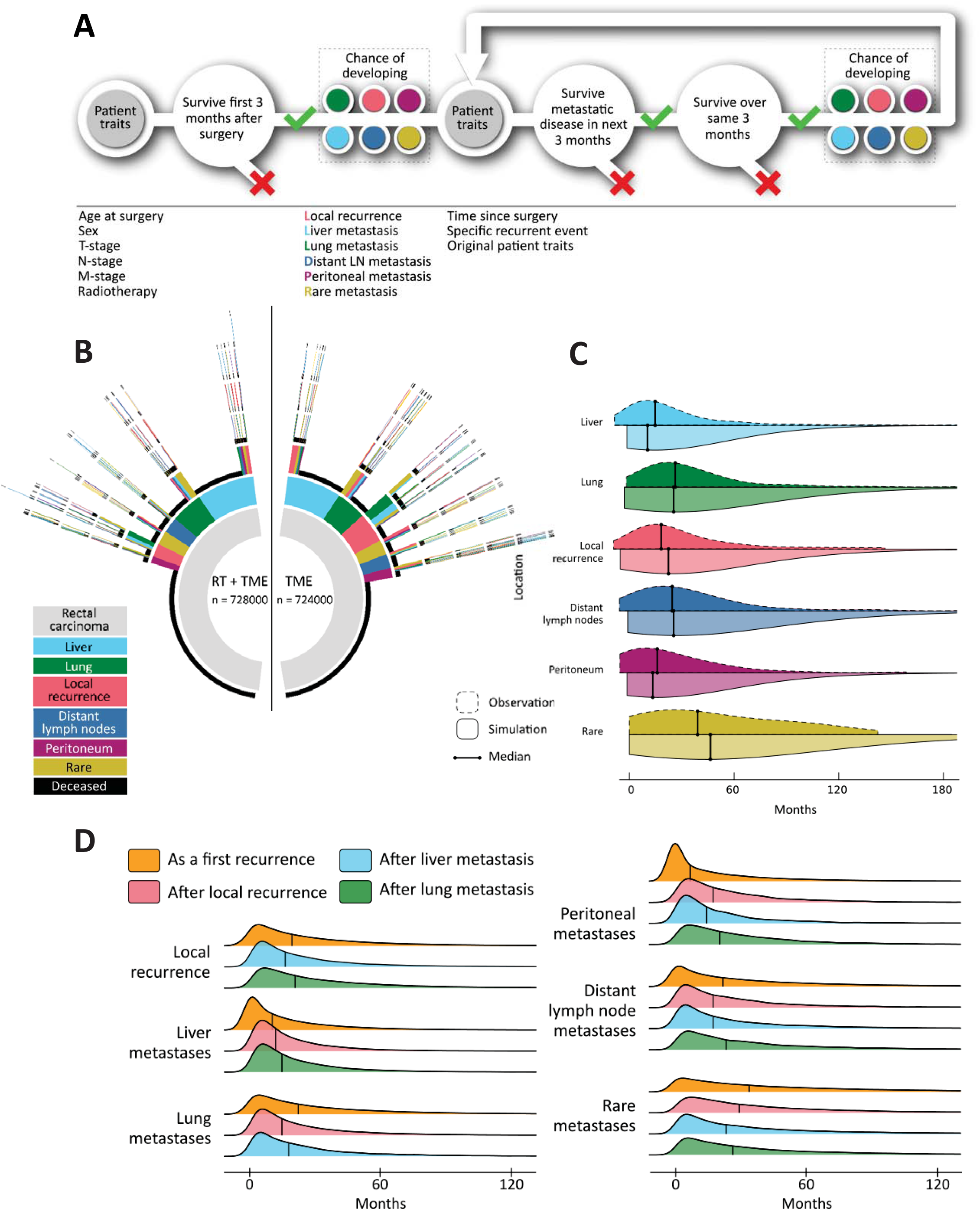
Metastatic patterns in the simulation model. A. **Flowchart** of the stochastic simulations, including decision points (survival) and probabilities of metastases developing at new locations (colored dots). B. Simplified circos plot of **metastatic sequences in the simulation** (n = 1,452,000), 566 different metastatic patterns were observed in the surgery only arm, in the radiotherapy arm 461 patterns were observed. C. **Timing of metastases** illustrated by split violin plot. Due to the increase in events and the longer follow-up period we observe a smoothing of the plot, but the median time to events in the simulated setting remains comparable with the observed setting. D. **Stacked density plot** based on the simulations: we determined the timing of subsequent events after 1) surgery (orange), 2) local recurrence (yellow), liver metastases (blue) and lung metastases (green).

### Metastatic spread simulated for individual patients

The second set of simulations of individual patients was used to study recurrence patterns systematically. For this purpose, we defined the median patient as a female of 65.8 years with a pT3 adenocarcinoma who was treated with surgery. The risks of developing subsequent recurrences strongly depended on the location of metastases, both synchronous and metachronous (Fig. 3a-f). The risks of local recurrence and liver and lung metastases were strongly reduced if metastatic spread (at another location) started 2 years after surgery compared to when it started synchronously (Fig. 3a-c). However, this difference was much less pronounced in the risks of distant nodal, peritoneal and rare metastases (Fig. 3d-f). Compared to the baseline (no synchronous metastases) the highest hazard ratios (HR) occurred for local recurrence when a synchronous metastasis was present at a rare location (HR 2.69, ranging 2.3-3.8 among patients varying in sex, treatment and initial lymph nodes), or for the risk of lung metastasis in the case of early local recurrence (HR 2.5, ranging 2.1-3.0). Hazard ratios were below 1 in some cases. For instance, in the presence of peritoneal metastases, the risk of liver metastases was strongly decreased (HR 0.10, ranging 0.08-0.135), probably due to strongly increased mortality rates. The simulated ‘median’ patients also differed systematically in sex, radiotherapy treatment and number of lymph nodes initially affected (0 or 3), but these factors had minor effects on resulting patterns in metastatic spread, death by metastatic disease or age at death. In figure 3 we therefore show simulation results averaged over the 8 combinations of these three binary variables.

**Figure 3.**
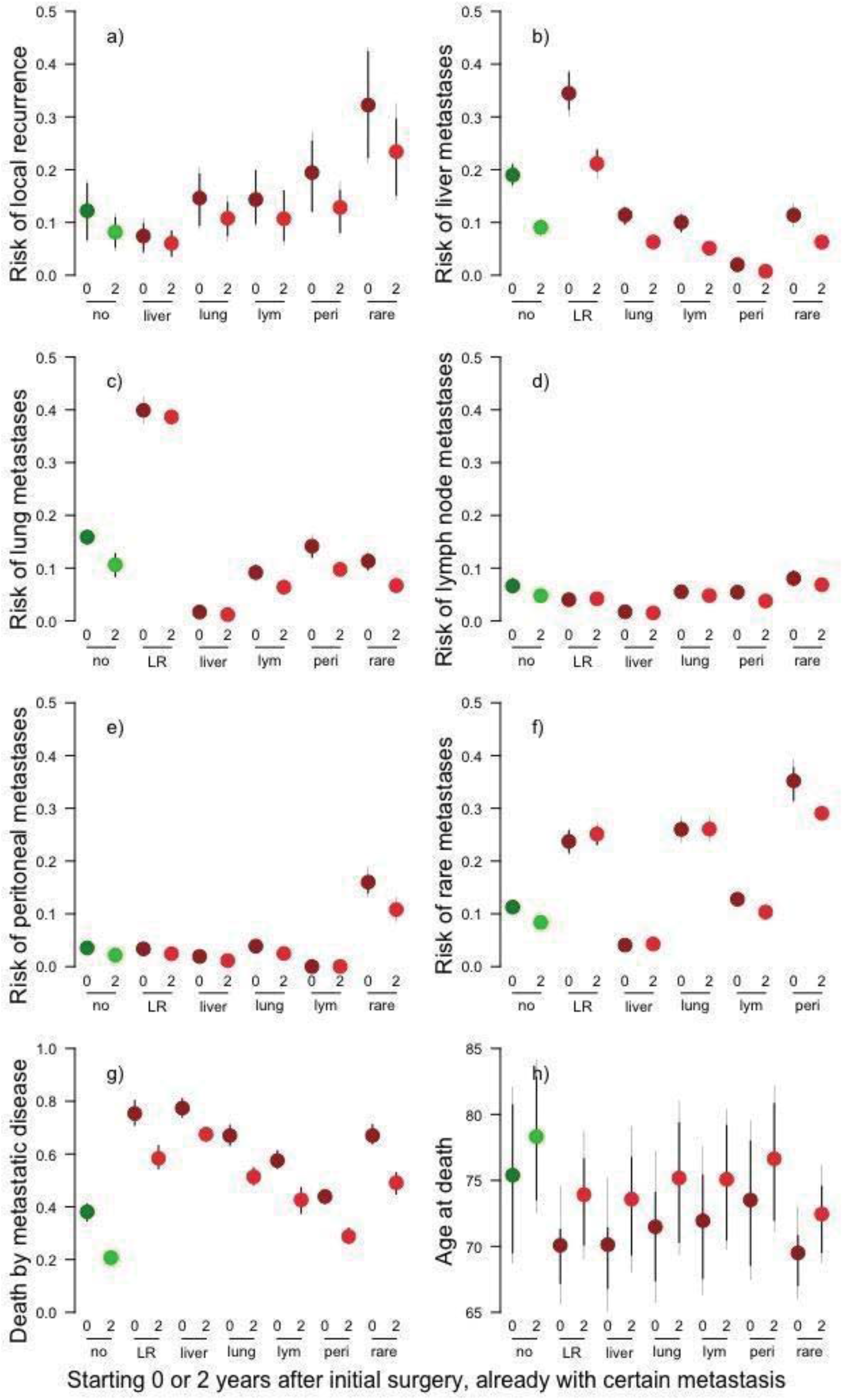
Results of simulations of ‘median’ patients. **a-f) Risk of the development of different types of recurrences,** depending on the initial traits: i. whether simulations started immediately after the initial surgery (t = 0, dark dot) or 2 years later (t = 2, light dot), and ii. whether a certain metastasis was already present at the start. **g) Proportion of simulations ending with death due to metastatic disease. h) Age at death of simulated patients**. Thin error bars indicate +/-1 sd, thick error bars indicate 50% of the variation. Only within-patient variation is used for the error bars. ‘no’: without recurrence at the start of the simulations, LR: local recurrence, lym: distant lymph node metastases, peri: peritoneal metastases.

### Risk assessment of metastatic cascades

In an attempt to evaluate the risks that specific metastases contribute to the development of subsequent new metastases at other locations, we performed additional regression analyses (details in Appendix) focusing on subsets of the trimesters which started with no or single-site recurrences. As proof of principle, for a female patient of average age (64.9 years old) with an average number of lymph nodes affected at the time of surgery (1.85) of a pT3 rectal adenocarcinoma and time set at one trimester after surgery, the predictions of these regression models are shown in figure 4. In those settings the presence of a local recurrence raised the chance of peritoneal metastases in the next trimester from 0.25% to 5.80%, for instance. Radiotherapy did reduce local recurrence risk from 1.30% to 0.47%, but when a local recurrence developed after radiotherapy, the risk of subsequent lung metastases was strongly increased (from 1.09% to 7.11%) (Fig. 4).

**Figure 4.**
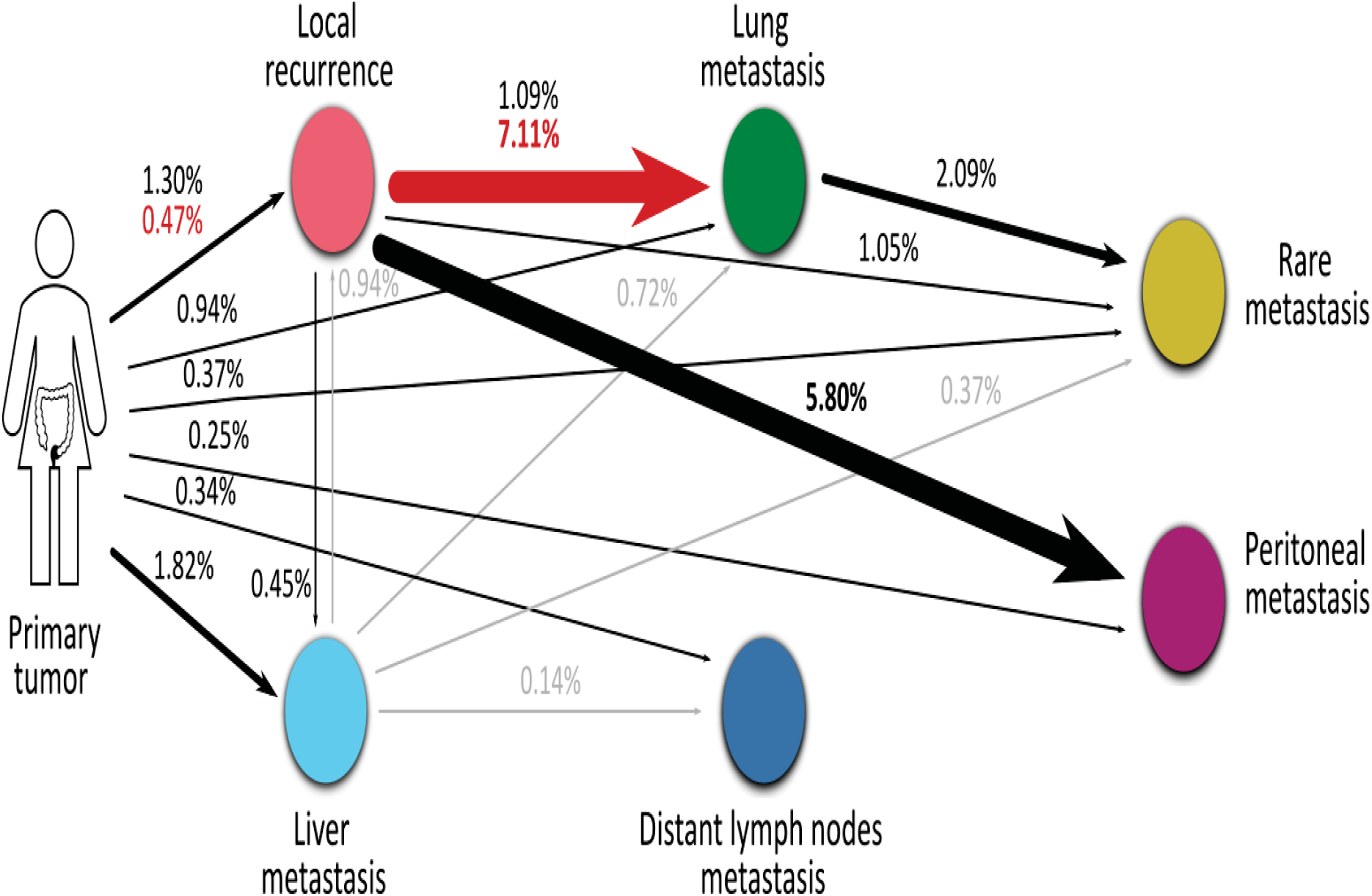
Risk assessment of metastases at new locations. Chance that within the next 3 months a recurrence is diagnosed at various locations given the starting conditions with no metastases or metastasis at single organs. Shown are model predictions for a female patient of average age (64.9 years old), average number of lymph nodes affected at the time of surgery (1.85) of a pT3 rectal adenocarcinoma, 3 months after initial surgery, not receiving radiotherapy. Fitted regression models were fitted with additive effects of these six factors. When radiotherapy had a significant effect, probabilities with radiotherapy are shown in red. Arrows with probabilities smaller than 0.01% are not shown. Thickness of the arrows indicate the 3-months risk.

## Discussion

There is a wide heterogeneity in disease progression in rectal cancer. With our new cross-disciplinary approach, we showed the variation and complexity of metastatic pattern analysis in a well-defined cohort of a randomized clinical trial for rectal cancer. In order to analyze this highly complex pattern of recurrences, including timing, location and different combinations, we applied methods from population ecology. Using a combination of nine logistic regression models we simulated a cohort of 1,452,000 rectal cancer patients, of which the combinations of metastatic locations, differences between the randomization arms (figure 2b) and timing of events (figure 2c) were comparable to the observation cohort. However, due to the size of this simulated cohort and due to the fact that the simulations were not right-censored like the observations, we were able to study combinations of metastases and timing of events in more detail.

Two types of simulation were presented in our paper. We simulated the whole cohort, which allowed us to study trends for large groups of patients, including rare metastatic cascades. All simulations ended with a mortality event (unlike the right-censored trial), giving the opportunity to study which metastatic combinations prove to be immediately fatal. In a second set of simulations, we studied the role of individual metastases more systematically, finding strong links between peritoneal and rare metastases (Fig. 3), for instance. When simulating the average female patient (figure 4) we were able to illustrate her relative risks of subsequent recurrences, thus describing a metastatic cascade based on statistical probabilities. Treatment with preoperative radiotherapy impacted the relative risk on recurrences. The risk of local recurrence was decreased after neoadjuvant radiotherapy, similar to findings in the observation cohort and other clinical trials(15, 17, 18). However, when local recurrences occur after radiotherapy, there is a selection of more aggressive tumors, which is reflected by a strongly increased risk of lung metastases in particular. This has also been observed in our clinical trial cohort(19).

Increasing knowledge on the development of recurrences is useful when discussing prognosis and personalizing surveillance for patients. Current surveillance strategies hardly take the individual situation into account and are mainly based on experience, rather than evidence(20, 21). Clinical trials that randomize between different surveillance strategies do not show differences in outcome(22, 23), and are not informative on timing of recurrences. Moreover, surveillance strategies are mainly aimed at the detection of the most common metastases. Risk assessment using the models we have developed (figure 3) allows for personalized surveillance, especially in patients after treatment for metastatic disease, based on starting point of surveillance and location of previous recurrences. This would facilitate cost-efficient strategies.

In addition to optimizing surveillance, it is also important to evaluate whether targeted interventions would change progression of disease. For this, we need detailed information on true patterns of metastatic spread. Are there metastatic cascades (metastasis-to-metastasis spread)(24) or are all metastatic clones derived early from the primary tumor in parallel progression(25)? In the latter case, the local microenvironment might be responsible for differences in growth velocity(26), which causes different times of presentation. If there is a possibility of metastasis-to-metastasis spread, then this would necessitate early interventions in small metastases. This view is supported by the good results in patients with liver metastasectomies(27, 28). Only when there are signs of local spread, in lymph vessels, the outcome is relatively poor(29). On the other hand, if all metastases are already in place, due to very early dissemination, there may be no benefit of resection of metastases. On the contrary, systemic treatment might be more effective at this stage. Very likely, multiple distinct routes of disease progression co-exist(30), with important clinical consequences. Information on these patterns is vital, since they determine the timing of interventions.

Once sufficient data on the actual origin and timing of metastases is available, a wide variety of transition models developed in population biology come within scope. Such data on cancer evolution will allow us to track the fate of individual clones and build models of recurrence that are characterized by tumor type(31), location and size (on a continuous scale, using Integral Projection Models(32, 33)). Tumor growth, colonization of metastatic location and resectability can thus be linked with histological features of the primary tumor, and with traits of the environment of the tumors, such as clinical characteristics of the patients. Impact of type and timing of systemic therapy can be modelled. Such recurrence models can then be used to study hypothesized mechanisms of metastatic spread in much more detail. Discrete stages during cancer progression, for example micrometastases, can be included in analogy with seed banks in invasive plant development(34) or dormant individuals in orchids(35).

When these models are combined with response functions that translate the size and distribution of the tumor cells into disease load and mortality risk of the patient, we can analyze how sensitive these output parameters are to changes in the model parameters. That means that we can use such population models to find out which parameters (e.g. growth of tumors at a certain location) contribute most to the discomfort of the patient. Furthermore, we could use those tumor population models to simulate which surveillance strategies (frequency, modality, target organs) are optimal when minimizing disease load as well as burden for patients(36).

The current study has several limitations, including the simplicity of the models and the used cohort. The current models are simplified, based on the input of a limited number of clinical data (figure 2a). However, they are adequate, given the similarities in metastatic patterns as well as the outcome for the average patient (figure 4). The addition of more information, in particular incorporating tumor biology would provide us with a higher accuracy. It is already known that tumor type(31) and molecular background(37, 38) influence metastatic patterns. The complexity of the model is limited by the number of cases that form the observation cohort. Incorporation of relatively uncommon features, such as tumor types other than adenocarcinoma n.o.s. or microsatellite instable rectal cancers(39) would allow more informative modelling. Larger well-defined cohorts are thus essential to increase accuracy. The observation cohort consists of rectal cancer patients, which automatically implies that our results cannot be directly translated into colon cancer, given their different metastatic patterns(40) and other treatment regimens. The cohort is based on patients operated between 1996 and 1999, which might be considered outdated by modern oncological standards. However, given the high level of quality control in particular to the quality of surgery(41) for the primary tumor, these patients can still be considered adequately treated. Their standardized and long follow-up is essential for studies of recurrent events.

When interpreting our simulation results it is important to keep in mind the order of events and the time step of the simulation (Fig. 2A). During the 3-monthly intervals we first simulated survival and only when a patient survived that time interval did we assess the establishment of tumors at new locations. This could lead to situations where new metastases are fatal immediately and thus not recorded. While peritoneal metastases were shown to be fatal within 3 months in more than 40% of the cases (Fig. 1A), our choice of 3-monthly censuses meant that such quickly developing diseases were statistically attributed to other factors that actually were present at the start of the interval. This ‘survivorship bias’(42, 43) is why the modeled effect of a peritoneal metastasis present at the beginning of an interval is not that detrimental (compare Fig. 3H with Fig. 1A). We chose this Markov chain approach (events during an interval are only affected by the state variables at the start of the interval) to keep our model as simple as possible, and used a time interval of 3 months to match the frequency of clinical examination during the trial. Obviously, future versions of the models can easily incorporate shorter time intervals or even continuous time if that would be relevant for specific research questions or for exploring different frequencies of patient check-ups.

In conclusion, we have confirmed the heterogeneity of recurrent rectal cancer, with the differential impact of recurrence locations on the outcome of patients. In addition, we have shown that methods derived from population dynamics can be used not only to describe these complex interactions, but even this relatively small, but well-described population can provide reliable information to predict outcome on individual patient level. Incorporation of additional (molecular) information will result in necessary insights in the progression of recurrent cancer.

## Data Availability

Data from the study are available on request

## Legends

**Supplemental table 1.** Clinical data of patients presenting with a rare metastasis as the first site of recurrence. These data are grouped in the category “other” in table 1.

| First site of recurrence |  |  |  |  |  |  |
| --- | --- | --- | --- | --- | --- | --- |
|  |  | Bone | Brain | Skin | Ovary | Bladder |
|  | Number of cases | 14 | 5 | 7 | 1 | 1 |
| age | Mean (years) | 67.9 | 65.0 | 60.0 | 55.0 | 60.0 |
| gender | Female | 0.8% | 0.4% | 0.4% | 0.2% | - |
|  | Male | 1.1% | 0.3% | 0.5% | - | 0.1% |
| treatment | RT | 1.2% | 0.5% | 0.3% | - | - |
|  | No RT | 0.7% | 0.1% | 0.7% | 0.1% | 0.1% |
| Distance from the anal verge | < 5 cm | 1.1% | 0.2% | 0.7% | - | 0.0% |
|  | 5-10 cm | 0.7% | 0.7% | 0.2% | - | 0.2% |
|  | 10-15 cm | 1.2% | 0.0% | 0.7% | 0.2% | - |
| TNM stage | I | 0.2% | - | 0.2% | 0.0% | 0.2% |
|  | II | 1.0% | 0.5% | 0.5% | 0.3% | - |
|  | III | 1.7% | 0.6% | 0.6% | - | - |
|  | IV | - | - | 1.1% | - | - |
| Tumor type | Adenocarcinoma | 0.9% | 0.3% | 0.4% | 0.1% | 0.1% |
|  | Mucinous ca | 1.3% | 0.7% | 0.7% | - | - |
| Number of recurrence locations | 1 | 76.9% | 60.0% | 71.4% | 100% | 100% |
|  | 2 | 23.1% | - | 28.6% | - | - |
|  | 3 | - | 40.0% | - | - | - |
|  | 4 | - | - | - | - | - |
|  | 5 | - | - | - | - | - |

## Appendix 1

Parameter values of the nine generalized linear regression models used for the stochastic simulations. The parameter values are the result of model-averaging (using the ‘full’ coefficients of the *model*.*avg* function of the R package *MuMIn*) of upto 4096 models (i.e. 212 perturbations of 12 explanatory variables). Continuous explanatory variables were normalized (mean=0, sd=1) prior to the logistic regression analyses. The 12 explanatory variables were *ageOp* (i.e. age at the time of TME surgery, mean=64.9, sd=11.1), *time* (i.e. number of trimesters since TME surgery, mean=20.5, sd=14.2), *depth* (i.e. invasion depth of the primary tumor (levels ‘pT3’ and ‘pT4’, with ‘pT1-2’ as baseline), *startLN* (i.e. number of regional lymph node metastases, mean=1.85, sd=3.92), *sex* (with Female as baseline), *treat* (treatment, with no radiotherapy as baseline), *lung* (whether there already has been a lung metastasis), *lymph* (whether there already has been a distant lymph node metastasis), *peri* (whether there already has been a peritoneal metastasis), *localR* (whether there already has been a local recurrence), *liver* (whether there already has been a liver metastasis) and *rare* (whether there already has been any of the other types of metastases). The nine binary response variables were: *diedFirst* = whether the patient died during the first trimester after surgery, and during following trimesters: *diedDis* whether a patient dies due to metastases during the next trimester, *diedNat* = whether a patient dies due to unrelated causes (conditional on surviving metastatic disease), *localRecur* = whether during the next trimester a local recurrence is discovered for the first time, and similarly for *newLiver, newLung, newLymph, newPeri* and *newRare*: whether metastases are discovered for the first time at each of these locations for the first time in the next trimester.

The mortality rate in the first trimester after surgery significantly increased with the age of the patient, and was higher for males. Later, the probability of mortality due to metastatic disease was increased by factors describing initial conditions (invasion depth, number of positive lymph nodes, patient’s age) and the presence of metastases at any of the locations (most of all liver, followed by rare locations), but decreased with time since surgery. Radiotherapy and sex did not significantly affect disease specific survival. Contrastingly, the chance of mortality due to other causes did increase with time (and patients’ age) and was higher in males. Of the metastasis-related variables, only the presence of a pT4 rectal cancer and the presence of rare metastases contributed significantly to background mortality.

The probabilities of new metastases appearing at various organs during a trimester generally followed the same patterns as described above (see details Appendix). Probabilities decreased with time since surgery and were higher with increasing invasion depth and higher lymph node stage. Local recurrence was decreased due to the application of preoperative radiotherapy, consistent with previous findings. In particular in the radiotherapy arm, the presence of local recurrences significantly increased the chance of liver, lung and rare metastases. Lung metastases were also more likely when liver metastases were already present. The probability of rare metastases significantly increased with the presence of other metastases except for liver metastases. Please note that these analyses of the probabilities of new metastases were conditional on the survival of a patient over the studied trimester. If, during the trimester, the patient died, additional metastases were not considered.

---

**appendix 1.**

|  | intercept | ageOp | time | depth3 | depth4 | startLN | sexM | treat | lung | lymph | peri | localR | liver | rare | anyMeta |
| --- | --- | --- | --- | --- | --- | --- | --- | --- | --- | --- | --- | --- | --- | --- | --- |
| diedFirst | -3.982 | 0.981 | 0 | 0.057 | 0.094 | -0.023 | 0.640 | 0.116 | 0 | 0 | 0 | 0 | 0 | 0 | 0.513 |
| diedDis | -6.343 | 0.315 | -0.683 | 0.832 | 1.372 | 0.228 | 0.110 | -0.020 | 1.284 | 1.145 | 0.414 | 0.893 | 2.075 | 1.453 | 0 |
| diedNat | -5.737 | 1.131 | 0.691 | 0.155 | 1.062 | 0.007 | 0.426 | 0.122 | -0.107 | 0.401 | 0.099 | 0.262 | 0.001 | 1.300 | 0 |
| localRecur | -6.935 | -0.006 | -0.913 | 1.337 | 1.202 | 0.300 | 0.008 | -0.864 | 0.327 | 0.479 | 0.407 | 0 | 0.133 | 1.607 | 0 |
| newLiver | -6.712 | -0.157 | -1.321 | 0.907 | 1.310 | 0.397 | 0.169 | -0.022 | -0.280 | -0.521 | -2.413 | 1.146 | 0 | -0.260 | 0 |
| newLung | -6.550 | -0.033 | -0.791 | 0.926 | 1.127 | 0.310 | 0.027 | 0.049 | 0 | -0.407 | -0.176 | 1.867 | -1.892 | -0.294 | 0 |
| newLymph | -6.923 | -0.014 | -0.647 | 0.572 | 0.455 | 0.497 | -0.069 | 0.008 | 0.189 | 0 | -0.230 | -0.011 | -0.752 | 0.807 | 0 |
| newPeri | -8.183 | -0.009 | -1.079 | 0.885 | 1.270 | 0.331 | -0.225 | -0.003 | 0.100 | -6.742 | 0 | 0.047 | -0.111 | 2.220 | 0 |
| newRare | -6.931 | -0.014 | -0.438 | 0.672 | 1.210 | 0.079 | -0.003 | 0.003 | 1.670 | 0.682 | 1.577 | 1.232 | 0.029 | 0 | 0 |

